# Efficiency and efficacy of sensing-monopolar recordings from the subthalamic nucleus in Parkinson’s disease at initial programming: a comparison to traditional methodology

**DOI:** 10.1101/2025.03.11.25323159

**Authors:** John A. Thompson, Erin M. Radcliffe, Steven Ojemann, Daniel R. Kramer, Pamela David-Gerecht, Michelle Case, Caleb Zarns, Abbey Holt-Becker, Robert S. Raike, Alexander J. Baumgartner, Drew S. Kern

## Abstract

**Background:** Optimizing deep brain stimulation (DBS) in Parkinson’s disease (PD) requires a complex process for evaluating clinical benefit and adverse effects. Localizing the contact with maximum beta (13-30Hz) power using a novel in-clinic sensing-monopolar (SenseMP) recording may improve the efficiency of selecting clinically optimal DBS settings.

**Objective:** Compare the clinical outcomes of optimal programming settings determined by SenseMP versus standard-of-care monopolar review (ReviewMP) for DBS therapy in PD.

**Methods:** Ten PD patients (4F/6M, post-diagnosis = 9.2 ± 3.5 years; on/off percent change = 43.7 ± 14.0) with bilateral subthalamic nucleus (STN) DBS were recruited. Independent evaluation of unilateral STN DBS was assessed with both SenseMP to identify the DBS contact detecting maximum beta peak power and ReviewMP. MDS-UPDRS Part III assessments for unilateral symptoms were compared between SenseMP and ReviewMP, and the time to complete these two conditions was recorded.

**Results:** Change from baseline in overall clinical outcomes did not differ between conditions (SenseMP = 60.5%, ReviewMP = 60.1% (p = 0.85); 19 hemispheres), nor were there differences in sub-analysis for bradykinesia, rigidity, and tremor. Using SenseMP to identify and conduct single contact ReivewMP was more time efficient than ReviewMP: 17.1 ± 4.2 minutes vs. 129.9 ± 24.9 minutes (p = 3.6e-15). SenseMP identified the maximum beta contact in SenseBP pairs at a specific level (17/20 times).

**Conclusion:** This study demonstrates that SenseMP localization of maximum beta peak power for DBS contact selection provides equivalent clinical efficacy to ReviewMP at a fraction of the required time.

## Introduction

Deep Brain Stimulation (DBS) of the subthalamic nucleus (STN) is a neurosurgical treatment option for Parkinson’s Disease (PD) complicated by motor fluctuations, bothersome dyskinesias, and/or pharmacological-resistant tremor^1–5^. Optimal DBS targeting and subsequent programming within the sensorimotor region of the STN effectively attenuates cardinal PD motor symptoms and enables reduced medication dosages^6–10^. Although recent advances in DBS electrode design have enabled independent contact control and directional field steering, the increased complexity and flexibility of these designs create multiple degrees of freedom, decreasing the efficiency of standard programming protocols^11–16^. Given the increased time demands and challenges presented by this expanding parameter space, efficient and objective strategies for DBS contact selection and programming, guided by neuroimaging, kinematic assessments, computational tools, and electrophysiological biomarkers, are under development and aimed toward delivering patient-centered therapy within a feasible clinical workflow. Several studies support the use of LFP features, specifically within the beta range (13-30 Hz), as potential biomarkers for guiding DBS programming in PD, but lack an intuitive process to select final stimulation contact due to sensing configurations available on implanted devices today^17,18^.

Current clinical-commercial tools permit spectral analysis of LFP features present in power spectral densities (PSDs). However, systematic strategies for quantifying and interpreting this data have not been standardized^19–21^. Available implementations use bipolar sensing (SenseBP) montages to determine the optimal DBS contact through meticulous comparison of maximal beta peak amplitudes present in LFP PSDs^19–21^. However, determining the largest peak amplitude differential between contact pairs is reliant on trial and error and requires expertise in interpreting computed measurements^12,18,19^. Furthermore, SenseBP configurations do not clarify which contact within or between a selected contact pair detects the maximal beta peak, complicating clinical interpretability.

In this study, we utilized an investigational Percept^TM^ SP Advanced Sense research system to enable a monopolar sensing (SenseMP) configuration for collecting LFP data. The contact segment that detected the greatest beta peak power based on SenseMP was then clinically programmed. Assessments of beta power measurements for each possible contact configuration (segment and level) were determined for SenseMP and for SenseBP pairs. Thereafter, standard-of-care monopolar review (ReviewMP) was performed. We then compared the time required and clinical outcomes of SenseMP versus ReviewMP.

## Methods

### Subject Information

Our study was carried out in accordance with the Colorado Multiple Institution Review Board (COMIRB # 21-4938) and the Declaration of Helsinki, with written informed consent obtained from all study participants. Patients were recruited from the University of Colorado Anschutz Medical Campus, Advanced Therapies in Movement Disorders program. All PD subjects underwent a standardized evaluation process for DBS candidacy and had at least a score of 2 or greater for any item assessing upper limb bradykinesia (items 3.4, 3.5, 3.6, 3.7, 3.8) and rigidity (item 3.3) of the Movement Disorder Society Unified Parkinson’s Disease Rating Scale (MDS-UPDRS) Part III in the off-medication state ^22^. Subsequently, subjects were implanted bilaterally into the STN with the Medtronic SenSight^TM^ B33005 directional DBS leads connected to the Percept^TM^ PC implantable neurostimulator with BrainSense^TM^. The B33005 DBS lead consists of four levels (termed 0, 1, 2, and 3) with 0.5mm vertical spacing, and the middle two levels are segmented into thirds (1a, 1b, 1c and 2a, 2b, 2c). A contact ring (i.e., 0, 3 or summation of a middle contact arrangement (e.g., 1a, 1b, and 1c)) is termed “level,” and an independent contact (e.g., 1a) is termed “segment”. The surgical procedure has been previously described^23^.

### Study Workflow

For each session, subjects withheld dopaminergic medication for at least 12 hours prior to the study visit. The following workflow was used for each session and is similar in design to previous work (Figure 1)^19^. In the -medication/OFF-stimulation condition (baseline) with the subject seated, eyes open and resting, a SenseMP recording was conducted for all levels (N=4) and segments (N=6) of the DBS lead. The investigational-use Percept^TM^ SP Advanced Sense research system was used to collect LFP data to enable the SenseMP configuration. This investigational system enabled collection of LFP data using the novel monopolar sensing configuration, the Percept™ SP Advanced Sense research system takes measurements between the electrode being sensed on the implanted DBS lead to a common reference electrode on the other implanted DBS lead. This enables isolation of activity from a single contact or set of contacts for SenseMP recordings. The implanted neurostimulator maintained standard commercial operation, and stimulation was turned off and not affected while using the research system. LFP data were collected from one hemisphere per session. A sensing montage that iterated through 5 predetermined configurations (SenseMP Segment, SenseMP Level, SenseBP Segment Set 1, SenseBP Segment Set 2, SenseBP Level) was performed to wirelessly stream LFP data at 250Hz for 55 seconds per recording. All recordings passed through two low-pass filters set to 100 Hz and two high-pass filters set to 1 Hz (for both bipolar and monopolar contact configurations). We collected all possible monopolar configuration recordings (levels = 4, segments = 6, total configurations = 10) and all possible bipolar configurations (n = 15; bipolar configurations are available on commercial systems). Three trials of each montage were collected to account for variability. The contact level and segment identity, maximum beta peak amplitude, and the frequency of the maximum beta peak were noted for each hemisphere. The contact with the maximum beta peak determined by SenseMP was used for comparison to the contact(s) identified by standard of care monopolar review (ReviewMP).

**Figure 1:**
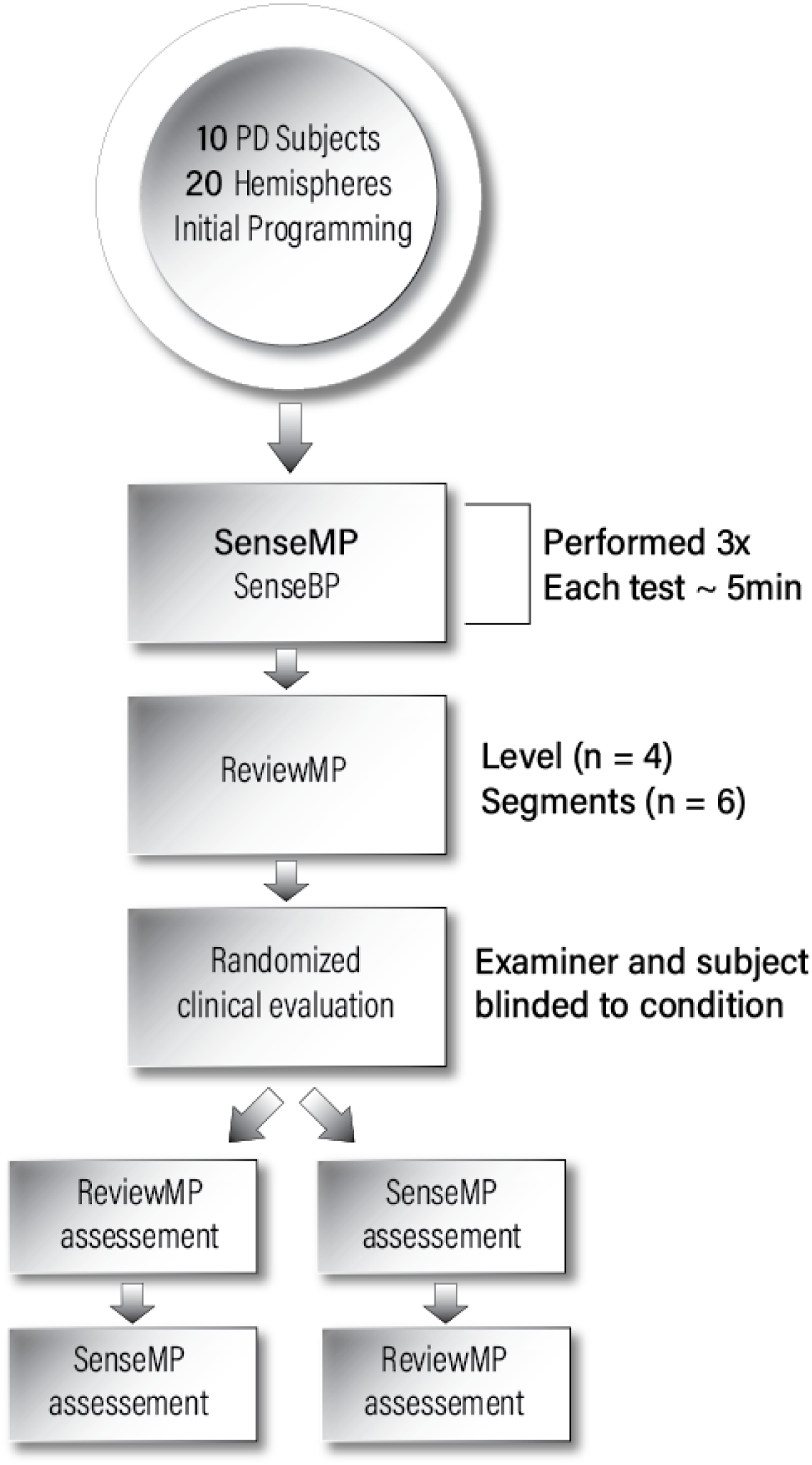
Workflow diagram of the study design

Subsequently, a traditional ReviewMP of all DBS levels (n=4) and segments (n=6) was performed by a movement disorders neurologist (examiner) blinded to the outcomes of the montage recordings^19^. Briefly, pulse width (60 μs) and frequency (125Hz) were maintained while amplitude was increased by 0.2 mA increments up to 5mA or until stimulation-induced adverse effects were observed. All side effects were recorded, including transient (those that resolved within 1 min) and persistent (those that lasted >1 min or included muscle contractions). Clinical benefit was determined by improvement in wrist and elbow rigidity. Upon completion, the clinician selected the contact(s), either the level or segment, that predominately had the greatest therapeutic window (TW), defined as the difference in DBS amplitude thresholds for initial clinical benefit and stimulation-induced adverse effects.

The total duration required to complete sensing (SenseMP and SenseBP) and ReviewMP were determined as follows: Recording of time was initiated for sensing once the research telemetry communicator was positioned over the neurostimulator and stopped upon verification of collection of all data at the conclusion of the third sensing research recording. For ReviewMP, recording of time was initiated upon placement of the clinical telemetry communicator and stopped at the end of testing the final contact. To determine the estimated time required to conduct the ReviewMP of the SenseMP identified contact, we used the following calculation. We estimated the total time per individual ReviewMP test (i.e., 0.2mA for 1ABC), by dividing the total time required for ReviewMP by the number of individual amplitude tests. Then, with the identified SenseMP contact, we computed the total duration for ReviewMP of that contact (i.e., all amplitude tests for 1A), by multiplying the individual ReviewMP test time by the number of amplitude tests for the identified contact. To this ReviewMP duration, we added the time required to evaluate SenseMP Level and SenseMP Segment (55 seconds x2). This value was compared to the total ReviewMP duration (Figure 2A).

**Figure 2:**
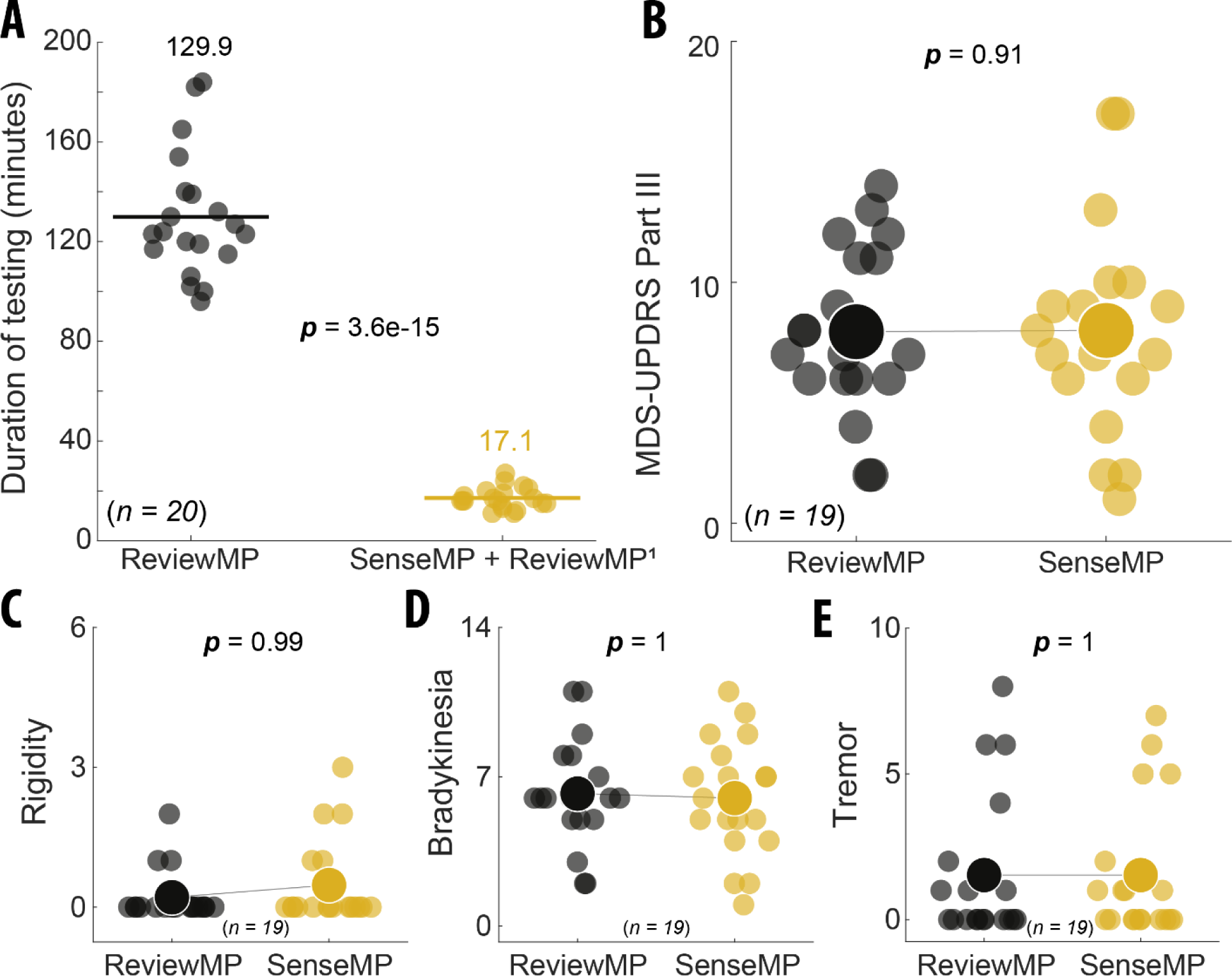
*A*) Total time (minutes) to complete ReviewMP (black circles) and sensing of monopolar and bipolar recording configurations to derive the contact with maximum beta power (SenseMP) in addition to ReviewMP of the identified contact (ReviewMP^1^) (gold circles). ReviewMP took on average seven times longer to perform than sensing. There was greater variability in duration to complete ReviewMP compared with SenseMP + ReviewMP^1^. Means are expressed for both conditions with horizontal lines. P-value is indicative of significant difference. *B*) Comparison of optimal contacts defined by ReviewMP (black circles) and SenseMP (gold circles), assessed in a double-blind comparison, resulted in equivalent motor symptom outcomes as measured by MDS-UPDRS Part III, in which each point represents a sum of the unilateral hemi-body sub scores of *C-E*. *C*) Unilateral hemi-body sub scores for rigidity (MDS-UPDRS items 3.3). *D*) Unilateral hemi-body sub scores for bradykinesia (MDS-UPDRS items 3.4, 3.5, 3.6, 3.7, and 3.8). *E*) Unilateral hemi-body sub scores for tremor (MDS-UPDRS items 3.15, 3.16, 3.17, and 3.18). Larger circles indicate the average for the sub scores as a function of the condition.

Clinical outcomes were compared between SenseMP and ReviewMP contact configurations based on random condition presentation and double blinding of the examiner and subject. Furthermore, the examiner was not present during the acclimation of each condition to minimize unblinding. The DBS amplitude for each condition was set at −0.2mA from the stimulation-induced adverse effect or up to 4.8mA based on ReviewMP assessment. Following a ∼2-minute wash-in phase, the examiner performed a unilateral MDS-UPDRS of the following items: wrist, elbow, ankle, and knee rigidity (item 3.3), bradykinesia (items 3.4, 3.5, 3.6, 3.7, 3.8), and tremor (items 3.15, 3.16, 3.17, and 3.18). Between conditions, stimulation was turned off, and the subject was assessed until a return to clinical baseline in the OFF-medication/OFF-stimulation state was confirmed by the examiner. Upon return to baseline, the examiner left the room to allow for acclimation to the second condition, and then the steps described above were then repeated. If the SenseMP and ReviewMP resulted in identical contact selection, then only one clinical assessment was performed, and the examiner was not notified until exiting the room. At the conclusion of both assessments, the blinded clinician selected the optimal configuration based upon clinical examination. To quantify the percent difference between each condition and the baseline OFF-medication / OFF-stimulation state at the beginning of the session, the following equation was used: (ABS(OFF-stimulation – ON-stimulation)/OFF-stimulation)*100. For the final clinical comparison, 19 out of 20 hemispheres were evaluated. For one hemisphere (4L), annotation error led to the incorrect selection of the contact with the maximum beta power for MDS-UPDRS comparison.

### Assessment of segment orientation

Medtronic SureTune™ 4 imaging software was used to perform DBS lead orientation analysis. Each subject had pre-operative anatomical imaging, including MRI T1 and T2 weighted, and a post-operative CT scan performed within 24 hours post-implant, as part of standard-of-care for DBS surgery. All images were imported into SureTune™ 4, and pre-operative MRI and post-operative CT images were fused to identify the anatomical location of the DBS lead. The Medtronic SenSight™ DBS lead has unique orientation markers that allow for automatic orientation of the lead enabled by SureTune™ 4 software. The anatomical direction for each segmented electrode was viewed from an axial slice and was recorded. The anatomical directions included lateral, medial, anterior, and posterior.

### Statistical Analysis

All statistical tests were conducted using MATLAB (v2024a). Mean differences in duration between SenseMP and ReviewMP were tested using a two-sided paired t-test (alpha = 0.05). Mean differences between MDS-UPDRS Part III total sub-scores were tested with an ANOVA, and individual sub-scores for rigidity, bradykinesia, and tremor were compared between conditions using Tukey’s HSD post-hoc pairwise comparisons. Level representation and segmented contact orientation differences between SenseMP and ReviewMP were compared using a Chi-Square test for independence. Within-subject hemispheric differences between SenseMP-determined maximum beta peak amplitudes and frequencies were compared with a two-sided paired t-test. All count comparisons for SenseMP-segment, SenseMP-level, SenseBP-segment, and SenseBP-level were conducted using Chi-square goodness of fit assuming equal distributions as the null hypothesis.

## Results

### Demographics

Ten PD subjects (females = 4 / males = 6; 69.0 ± 4.5 years old at time of study; disease age of onset: 61.4 ± 6.7; PD duration in years: 9.2 ± 3.5; mean ± standard deviation – throughout the text, ‘±’ will refer to the same descriptive statistics) participated in the study. Table 1 provides the demographics of the subjects. Sessions were conducted following lead implantation surgery 18.2 ± 7.3 days post-implantable neurostimulator implant and 27.2 ± 6.3 days post-lead implant. Across subjects, sessions were separated by an average of 2.5 ± 2.5 days. Analysis was performed for both right and left hemispheres per subject for a total of 20 hemispheres.

**Table 1:**
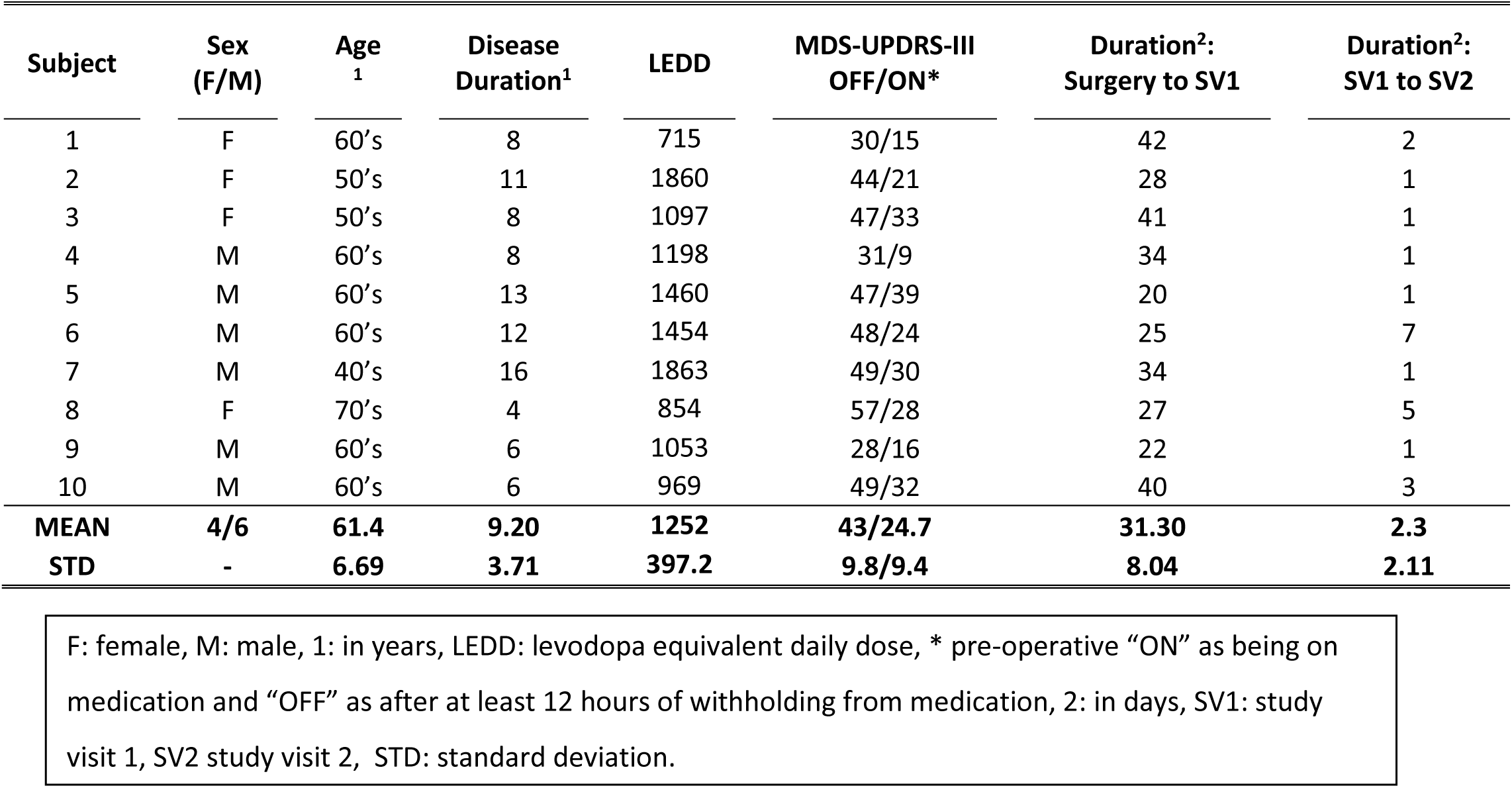
Subject Demographics.

### Within-subject comparisons of SenseMP beta parameters

We previously described the variability of beta peak frequency and amplitude using the same platform, excluding SenseMP^18^. Therefore, in the current study, we focused on SenseMP to evaluate within-subject differences across trials. There was no difference in within-subject beta peak frequency, amplitude, or contact determined by maximum beta power across three trials (peak: F(2,57) = 0.155, p = 0.85; amplitude: F(2,57) = 0.016, p = 0.98; contact: F(2,57) = 0.063, p = 0.93). Analysis was therefore performed on resulting PSDs resulting from the average across the three recording samples.

Table 2 denotes subject and hemisphere-based observations of the beta peak frequency and power identified by SenseMP and the corresponding beta peak power of ReviewMP. No differences were observed between left and right hemispheres for the peak frequency at which the maximum beta power was derived (right Hz = 16.2 ± 2.22, left Hz = 16.9 ± 2.59; *t*(9) = 1.07, *p* = 0.3), or for the amplitude of the maximum beta microvolt peak (right µVp = 3.15 ± 2.46, left µVp = 3.09 ± 1.68); *t*(9) = −0.101, *p* = 0.92). The contact with the second-greatest beta peak was on the same level as the contact with the greatest beta peak in 17/20 cases. As an example, if 1c had the highest beta peak power measured on the segments, then the 1a or 1b segment had the second-highest beta peak. In 17/20 hemispheres, the SenseMP defined segment was on the level of the maximum beta peak. In the three hemispheres that differed, the level of the maximum beta peak was on the most ventral (2) or dorsal contact (1), which are not segmented. In these three cases, the SenseMP level with the second highest beta peak power was a segmented level and corresponded with the SenseMP segmented contact. In 19/20 hemispheres, SenseMP resulted in a segment contact. In all 19 cases that resulted in a segment contact, the SenseMP segment corresponded with the SenseMP level of segments (1abc or 2abc) that had the greatest beta power peak of only the segmented levels.

**Table 2:**
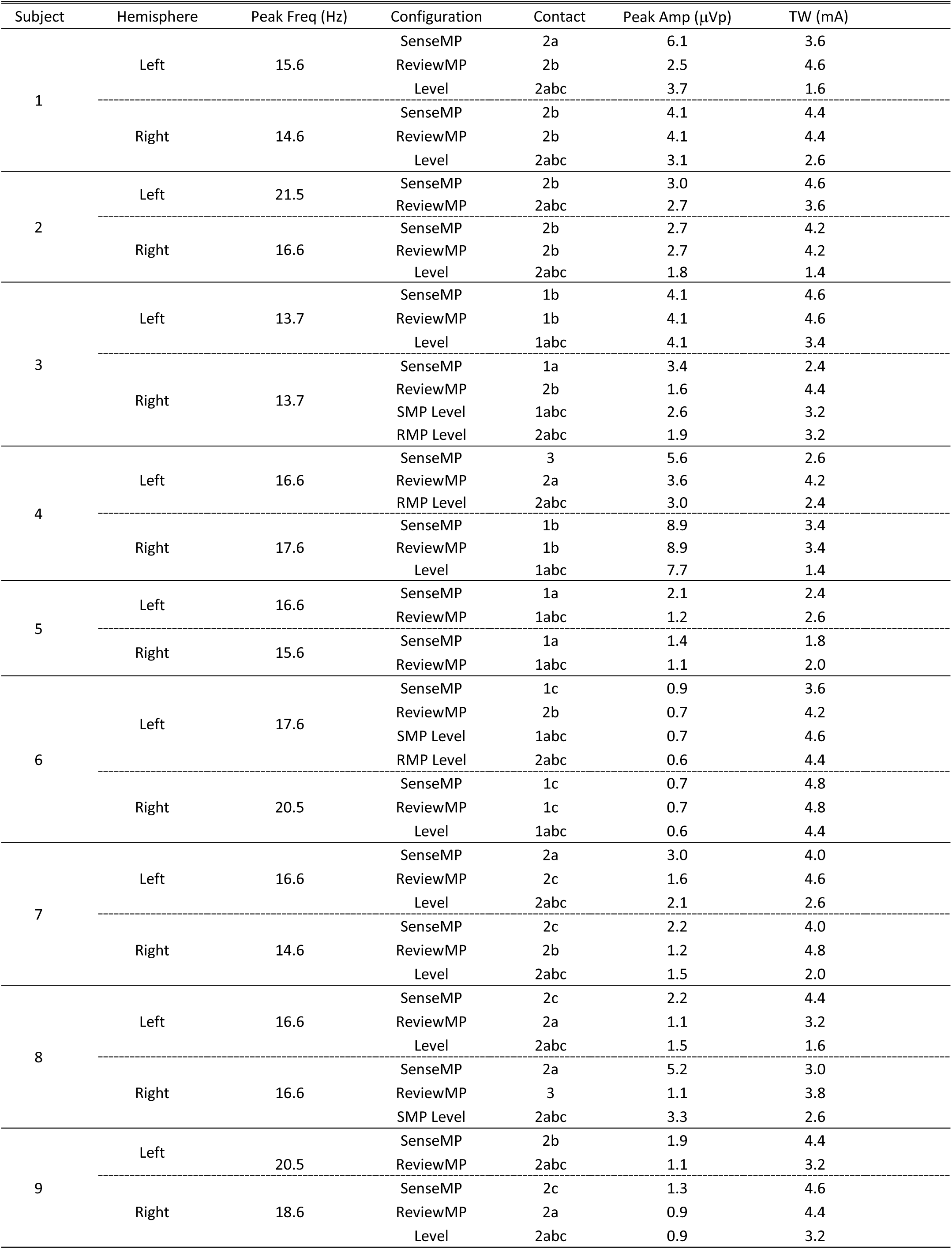
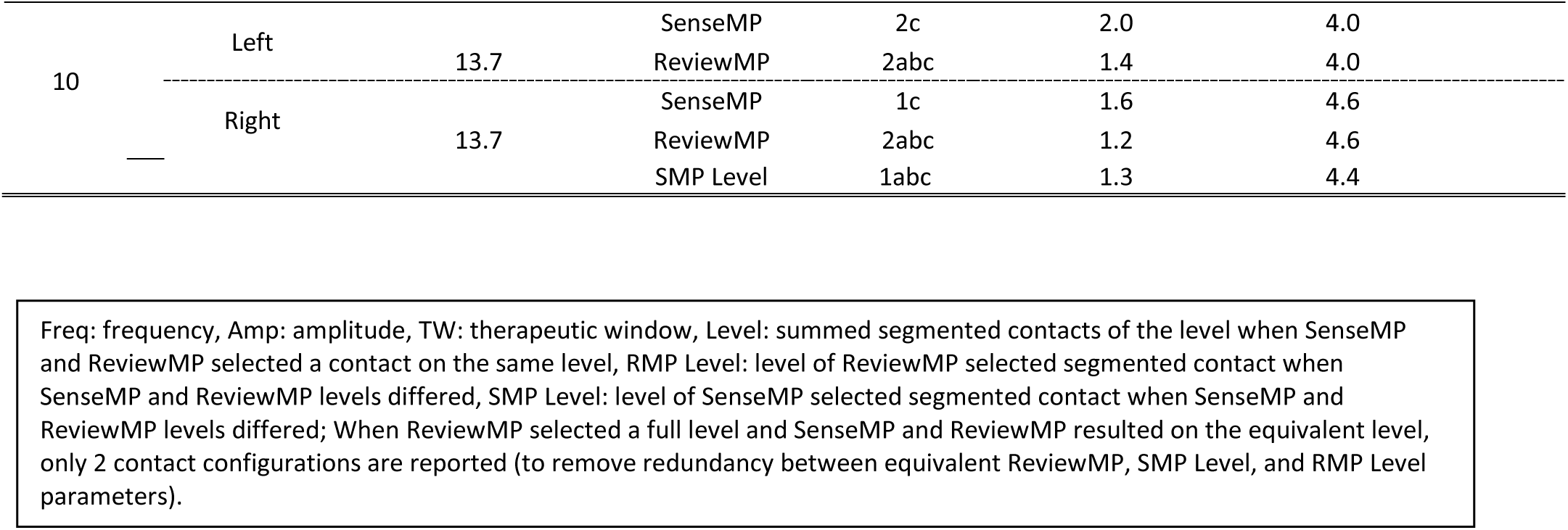
SenseMP and ReviewMP of lead segment and level.

### Efficiency differences between SenseMP and ReviewMP

To represent utilization of SenseMP in clinical practice, the duration to complete ReviewMP of the SenseMP identified contact(s) with maximal peak beta power (ReviewMP^1^) was compared to the duration of ReviewMP. Across subjects, the duration of SenseMP (single recording for both monopolar configurations; levels and segments) combined with the ReviewMP of the SenseMP identified contact was 17.1 ± 4.2 minutes. The total average duration for the ReviewMP was 129.9 ± 24.9 minutes across all patients. These durations were significantly different between conditions (*t*(19) = 22.53, p = 3.59e-15; Figure 2A). The duration to complete ReviewMP was of greater variability compared with sensing across all subjects, as evident in the differences in standard deviation.

### Motor outcome measures between SenseMP and ReviewMP

Combined MDS-UPDRS sub-scores for rigidity, bradykinesia, and tremor were not significantly different between conditions (*t*(18) = −0.10, *p* = 0.91). A complementary analysis was conducted on the percent difference values derived for each condition compared to baseline using a paired t-test (*t*(18) = 0.185, *p* = 0.85; mean percent difference SenseMP = 60.5 ± 15.3 and ReviewMP = 60.1 ± 13.5). In addition, an ANOVA for condition by sub-score (Figure 2B) demonstrated a non-significant omnibus for condition (*F*(1,108) = 0.002, *p* = 0.96) and condition by sub-scale interaction (*F*(2,108) = 0.12, *p* = 0.887). Post-hoc Tukey HSD comparisons for the sub-scores demonstrated no differences between conditions (rigidity: *t*(108) = −0.38, *p* = 0.99 (Figure 2C); bradykinesia: *t*(108) = 0.308, *p* = 1 (Figure 2D); tremor: *t*(108) = 1.43e-15, *p* = 1 (Figure 2E)).

### Comparison of therapeutic window between SenseMP and ReviewMP

ANOVA comparison of the SenseMP Level, SenseMP Segment and ReviewMP indicated a significant difference in the TW (*F*(2,56)=9.48, *p* < 0.001). Post-hoc Tukey’s HSD tests indicated that TW for ReviewMP (3.98 ± 0.76mA) and SenseMP of the segment (3.83 ± 0.87mA) were not significantly different (*t*(56) = 0.52, *p* = 0.86). However, both SenseMP of the segment and ReviewMP had greater TW than SenseMP of the level (2.85 ± 1.01mA; *t*(56) = −3.4, *p* = 0.003 and *t*(56) = 4.01, *p* < 0.001, respectively; Table 2). In three of the twenty hemispheres (2L, 8L, 9L), the contact selected by ReviewMP differed from the contact with the greatest TW, due to the selected contact having a lower stimulation amplitude to eliminate upper extremity rigidity. In one hemisphere (9R), the contact selected by ReviewMP was selected because it induced dyskinesias at higher stimulation amplitudes.

### Comparison of SenseMP and ReviewMP in contact selection and therapeutic respons

Qualitative assessment and comparison of contact level localization between SenseMP and ReviewMP demonstrated that for 15/20 hemispheres, the condition-derived optimal contact configurations were on the same level (Figure 3A). In 5/20 hemispheres, the SenseMP and ReviewMP selected segments were identical. In 7/20 hemispheres, ReviewMP selected a level, while SenseMP only selected a level in 1/20 hemispheres,. Quantitative analysis demonstrated no difference between SenseMP and ReviewMP for level localization of optimal contact configurations (χ^2^(3) = 5.33, *p* = 0.37). For the 5 hemispheres determined to be on different levels between conditions, the ReviewMP derived contact was superior to the SenseMP derived contact level in 4/5 cases. Orientation analysis was conducted on the 8 hemispheres with ReviewMP selecting segmented contacts that could be directly compared to the SenseMP segments. Qualitative assessment (Figure 3B) demonstrated an apparent lateral bias for segments selected by ReviewMP and, conversely, a medial bias for segments selected by SenseMP. However, quantitatively this difference was non-significant (χ^2^(7) = 9.3, *p* = 0.15).

**Figure 3:**
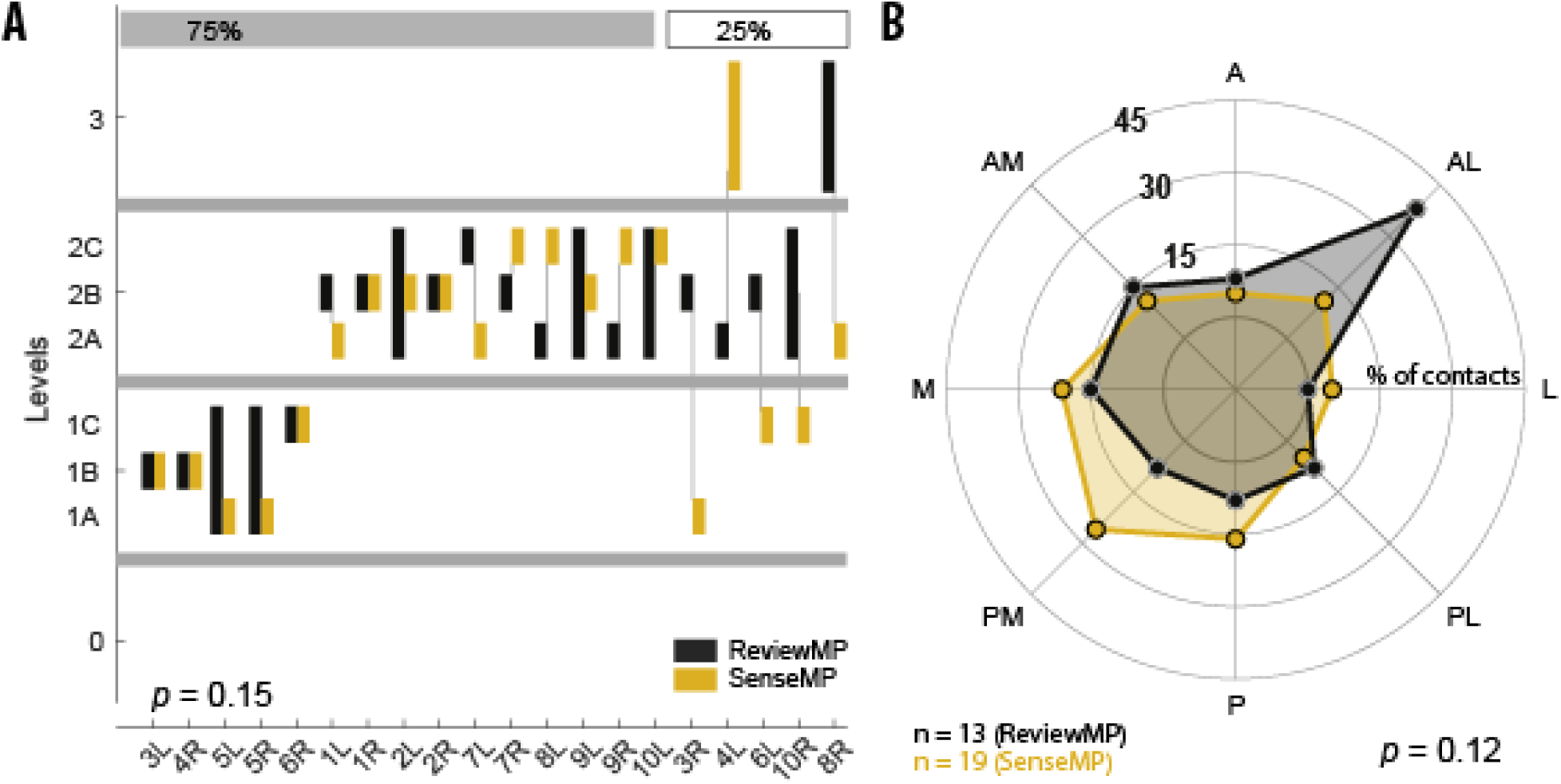
*A*) In 15/20 hemispheres, the SenseMP and ReviewMP contacts were localized within the same level, and for 5/20 hemispheres the SenseMP and ReviewMP contacts were localized one level apart. *B*) The distribution of segment orientations for SenseMP contacts exhibited a greater proportion in the medial region of STN, whereas the ReviewMP segment contacts appeared to have a greater proportion in the lateral region of STN. A: anterior, AL: anterior-lateral, AM: anterior-medial, L: lateral, M: medial, P: posterior, PL: posterior-lateral, PM: posterior-medial.

Visual inspection demonstrated more easily differentiated beta peaks with SenseMP compared with SenseBP for level (Figure 4A & 4B), which was even more apparent for segments (Figure 4C & 4D). Statistical analysis of the observed distribution for SenseBP level configuration based on maximum beta power (i.e., 0-level between contacts, 1-l evel between contacts, and 2-level between contacts) demonstrated a significant difference in relative count (with respect to the expectation of equal representation; χ^2^(2) = 6.1, p = 0.047; 0-level = 7, 1-level = 11 and 2-level = 2) (Figure 4E & 4F). We next evaluated the alignment of the SenseMP maximum beta level with the SenseBP configuration, with respect to whether SenseMP level was aligned with the superior (top), inferior (bottom), or middle contacts of the SenseBP configuration. The analysis of the observed distribution of SenseMP for level identified maximum beta SenseBP contact did not result in a significant difference in relative count (with respect to the expectation of equal representation; χ^2^(2) = 4.9, p = 0.086; Superior = 11, Inferior = 6, and Middle = 3). Notably, for 85% of the comparisons, the maximum SenseMP level contact was one of the SenseBP contacts (17/20), with the remaining three SenseMP level contacts identified as the ‘middle’ contact of a 1-level or 2-level between contact SenseBP configuration (Figure 4E & 4G).

**Figure 4:**
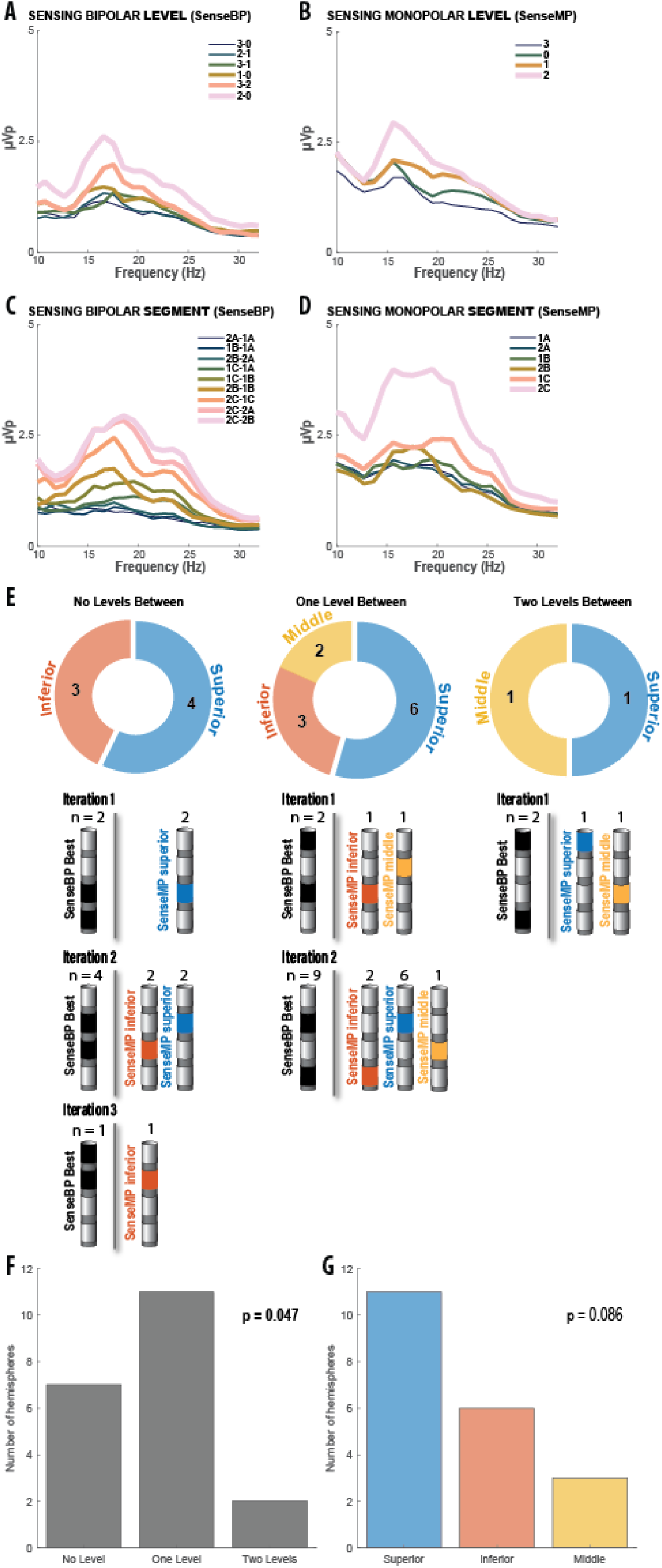
Comparison of SenseMP and SenseBP configurations. *A-D*) A single representative set of LFP recordings from a single subject’s hemisphere. Power spectral density plots depict differences in power within the beta frequency range. *A*) SenseBP for contact levels. *B*) SenseMP for contact levels. *C*) SenseBP for segments. *D*) SenseMP for segments. *E*) Schematic representation and summary of contact configurations derived from the SenseBP levels with the highest beta power paired with their corresponding SenseMP level with the highest beta power. *F*) Number of hemispheres exhibiting the three possible bipolar configurations: No Level (0-level between contacts), One Level (1-level between contacts), and Two Levels (2-level between contacts). *G*) Number of hemispheres exhibiting the three possible corresponding positions of SenseMP level contacts with respect to the SenseBP configuration.

In an analysis of the three subject hemispheres identifying the SenseMP contact as between the two SenseBP contacts, the difference in peak beta power between the maximum SenseMP contact and a subsequent contact of SenseBP was minimal. For Subject 2L (i.e., Subject 2, Left STN), the peak beta power of the SenseMP was 2.7μVp, and the second-best contact, as identified by beta, had a peak beta power of 2.2μVp. Subject 6, for both left and right hemispheres, had minimal difference (<0.1μVp difference), as assessed by SenseMP beta peaks between two contacts, with the second highest peak contact being identified by SenseBP.

Segmental analysis indicated that the contacts of SenseBP consistently identified one of the SenseMP contacts (20/20; χ^2^(1) = 20, p = 7.74e-6). In 12/20 cases, the SenseMP segment was the superior contact; in 6/20 cases, it was the inferior contact of the two SenseBP contacts (χ^2^(1) = 2, p = 0.15). Notably, only 2/20 cases of the bipolar configuration were on the same level (e.g. 2a-2b) with all other 18/20 cases being on adjacent levels but on the same vertical segment orientation (e.g. 1a-2a; χ^2^(1) = 12.8, p = 3.46e-4). In no case was there a deviation of SenseBP being on different levels and of different segmental orientations (e.g. 1a-2c). Furthermore, in no cases was the SenseMP segment between SenseBP contacts within the same level.

## Discussion

In this study, we compared the efficiency and efficacy of using maximum beta power recorded during SenseMP versus traditional ReviewMP to select the optimal therapeutic DBS contact in PD subjects undergoing initial programming with STN-DBS. The key findings from this study were 1) no significant difference in clinical outcome, as measured by MDS-UPDRS Part III (Items 3.4, 3.5, 3.6, 3.7, 3.8, 3.15, 3.16, 3.17, and 3.18) between SenseMP and ReviewMP, 2) non-inferiority at time of initial programming of SenseMP for determination of initial DBS contact selection, 3) an 85% shorter duration in obtaining contact selection using SenseMP as compared to ReviewMP, 4) 75% agreement for contact level selection between SenseMP and ReviewMP, 5) greater qualitative differentiation between contacts based on beta power magnitude for SenseMP than SenseBP (commercially available sensing configuration), 6) 85% agreement between SenseMP identified contact levels and one of the SenseBP contact levels.

Results from this study are well positioned within growing literature on the potential utility of commercially accessible LFP montages in DBS programming based upon beta power^18–20^. Recently, in a randomized, blinded, three-condition trial in STN-DBS treated PD subjects, the clinical efficacy and time efficiency of ReviewMP were compared to both imaging-guided and SenseBP-guided programming^17^, finding no difference in clinical efficacy but significantly improved efficiency for both imaging-guided and SenseBP-guided compared to ReviewMP-guided programming. In our study, the time duration to complete the SenseMP condition included recordings for both SenseMP and SenseBP, repeated three times. Clinical efficiency is also impacted by the capacity to differentiate between contacts based on maximum beta power. In this regard, SenseMP qualitatively permits more clearly resolved differences between contacts based on beta power, when compared to SenseBP, and does not require the use of complex algorithms or real-time problem solving to determine the optimal contact within the bipolar pair to program^18^. As exemplified by Binder et al., (2023)^17^, there was ambiguity in which contact to stimulate within SenseBP-identified pairs, resulting in the group electing to evenly distribute stimulation between the two segmented contacts in the pair. The regional bias appreciated in our study may corroborate recent reports validating that bradykinesia suppression appears to result from more medial modulation of the STN, and beta power is known to correlate more prominently with bradykinesia^24^. There is also evidence that rigidity suppression may have a superior bias in the STN, which may partially explain our observation that in 5/20 hemispheres, the ReviewMP optimal contact was more superior to the contact selected by SenseMP^25^. Finally, the TW of SenseMP of the level, but not of the segment, was significantly less compared with ReviewMP. In five conditions, the SenseMP level was more ventral to ReviewMP contact(s) and had a lower threshold to induce dyskinesias (N=2), paresthesias (N=2) and hyphophonia (N=1).

In this first use of SenseMP with fully implanted systems, we focused on identifying the contact with maximum beta power, but other neurophysiological markers other than beta may be of great value (e.g. alpha, gamma) and should also be studied, especially considering the heterogeneity of PD symptoms. SenseMP is a potentially powerful tool in DBS programming; however, it currently cannot be employed in patients with 1) unilateral implants, 2) bilateral lead implants driven by separate neurostimulators, or 3) implanted systems with high impedance. Consequently, the use of SenseBP will be of value in these scenarios and as our study suggests, one of the contact pairs having the greatest peak beta may be selected for programming. Finally, SenseMP resulted in a predominant bias to select segmented contacts (19/20) due to the greater surface area of the contact as well as the surgical targeting strategy in the current study.

There are several limitations to our study, though many are inherent and acknowledged in clinical practice. For example, ReviewMP is highly operator-dependent and potentially prone to bias, as evident in our study. In addition, a selected contact may have greater improvement of motor symptoms at lower stimulation but also have a lower amplitude threshold to induce adverse effects compared to an adjacent contact. Such a contact may have a lower TW but be clinically more ideal since greater motor response is achieved with less stimulation^26^. Furthermore, although the MDS-UPDRS is an established and validated scale, subtle differences in therapeutic response across tested contacts may not be adequately represented or appreciated by the examiner. Another limitation of the study is that evaluations were only performed in the acute setting and the long-term effects of programming by SenseMP are unknown.

This study demonstrates that SenseMP STN DBS programming may be a time-efficient and clinically effective tool in DBS programming for PD patients. It is likely that SenseMP will allow practitioners to focus on testing contacts with ReviewMP that have the greatest therapeutic benefit, thus better utilizing clinical time. It will be important to evaluate potential future research endeavors and clinical application of SenseMP as this technology becomes more widely available.

## Data Availability

All data produced in the present study are available upon reasonable request to the authors

## ACKNOWLEDGEMENTS

We are grateful for the patients who participated in this study. Funding for this investigator-initiated study was provided by Medtronic.

## AUTHOR CONTRIBUTIONS

EMR: collected data, drafted original version, revised final draft; DRK: revised final draft; SO: collected data; MC: contributed analysis tools, revised paper; CZ: contributed analysis tools, revised paper; AHB: contributed analysis tools, revised paper; RR: contributed analysis tools, revised paper; AJB: collected data; DSK: conceived of study, collected data, drafted original version, revised final draft; JAT: conceived of study, collected data, developed analysis tools, drafted original version, revised final draft.

